# Does Smoking Cause Lower Educational Attainment and General Cognitive Ability? Triangulation of causal evidence using multiple study designs

**DOI:** 10.1101/19009365

**Authors:** Suzanne H. Gage, Hannah Sallis, Glenda Lassi, Robyn Wootton, Claire Mokrysz, George Davey Smith, Marcus R. Munafò

**Author notes:** Corresponding author: Suzanne Gage; Department of Psychological Sciences, University of Liverpool, Eleanor Rathbone Building, Bedford Street South, Liverpool, L69 7ZA, UK.

## Abstract

**Objectives:** Observational epidemiological studies have found associations between smoking and both poorer cognitive ability and lower educational attainment; however, evaluating causality is more challenging. We used two complementary methods to attempt to ascertain whether smoking causes poorer cognitive ability and lower educational attainment.

**Design:** A cohort study (Study One) and a two-sample Mendelian randomization study using publicly-available summary statistics (Study Two).

**Setting:** The Avon Longitudinal Study of Parents and Children (ALSPAC), a birth-cohort study based in Bristol, United Kingdom, and general population samples from published genome-wide association studies (GWAS).

**Participants:** Up to 12,004 young people in ALSPAC (complete case analysis N = 2,107) (Study One and Study Two), and summary statistics from three previously published GWAS (not individual-level data) (Study Two).

**Main outcome measures:** Cognitive ability at age 15 (assessed via the Wechsler Abbreviated Scale of Intelligence) and educational attainment at age 16 (assessed via school records) (Study One), and educational attainment (measured as years in education) and fluid intelligence from previously published GWAS (Study Two).

**Results:** In Study One, heaviness of smoking at age 15 was associated with lower cognitive ability at age 15 and lower educational attainment at age 16. Adjustment for potential confounders and earlier cognitive ability or educational attainment attenuated findings although evidence of an association remained (e.g., fully adjusted cognitive ability beta - 0.736, 95% CI −1.238 to −0.233, P = 0.004; fully adjusted educational attainment beta −1.254, 95% CI −1.597 to −0.911, P < 0.001). Comparable results were found in sensitivity analyses of multiply imputed data. In Study Two, two-sample Mendelian randomization indicated that both smoking initiation and lifetime smoking lower educational attainment and cognitive ability (e.g., smoking initiation to educational attainment inverse-variance weighted MR beta −0.197, 95% CI −0.223, −0.171, P = 1.78 × 10^−49^). Educational attainment results were robust to various sensitivity analyses, while cognition analyses were less so.

**Conclusions:** Our results provide evidence consistent with a causal effect of smoking on lower educational attainment, although were less consistent for cognitive ability. The triangulation of evidence from observational and Mendelian randomisation methods is an important strength for causal inference.

**Summary boxes:** *What is already known on this topic:* Associations are seen between smoking and both educational attainment and cognition. These is some evidence that educational attainment might causally influence smoking, but causality in the opposite direction has not been assessed.

*What this study adds:* Using multiple methodologies, we found evidence consistent with a causal effect of smoking on lower educational attainment. An exploration of potential mechanisms could inform the development of interventions to mitigate this risk.

## Introduction

Cigarette smoking is associated with numerous adverse health outcomes. Smoking prevalence has declined in high income countries in recent years, but this decline has been strongly socially patterned, with the greatest declines in the most advantaged sections of society (1, 2). As a result, smoking is now a major driver of health inequalities between advantaged and disadvantaged socioeconomic groups. There is evidence that educational attainment causally influences the likelihood of smoking initiation, and also heaviness of smoking and likelihood of quitting among smokers (3, 4). However, it is less clear whether smoking has an impact on educational attainment.

There is some evidence that young people with a history of smoking show poorer academic and occupational outcomes compared to individuals who do not smoke or desist soon after a period of experimentation (5-8). More generally, substance use in adolescence has been reported to be associated with poorer educational attainment (9-11). However, it is possible that the observed negative association between substance use and both cognitive ability and educational attainment may result from students experiencing academic failure being more likely to engage in substance use, for example in an attempt to manage resulting negative feelings (12-14). In this study, we assessed the association between cigarette smoking and cognitive ability and educational attainment in adolescents. We hypothesised that smoking cigarettes would be associated with lower cognitive and educational outcomes.

We first used data from a large UK based pregnancy cohort, the Avon Longitudinal Study of Parents and Children (ALSPAC), to investigate whether smoking, measured at age 15, was associated with cognitive ability, assessed at the same age, and educational attainment at age 16, before and after adjustment for a range of potential confounders. We attempted to minimise the impact of reverse causation by adjusting for pre-existing general cognitive ability (in the general cognitive ability analyses) or earlier educational attainment (in the education analyses). In an attempt to account for selection bias due to the typical attrition of large cohorts, as a sensitivity analysis we imputed up to 100 datasets and repeated the analyses.

We next used Mendelian randomization analyses of publicly-available summary data to confirm these findings in an independent sample. Mendelian randomization (MR) uses genetic variants as proxies for the exposure of interest to overcome problems of residual confounding and reverse causality (15, 16), allowing for stronger causal inference. This use of multiple methods, each with different and ideally uncorrelated strengths, limitations, and sources of bias allows the triangulation of results to support more robust causal inference (17-19).

## Study One

### Methods

#### Data sources

The Avon Longitudinal Study of Parents and Children (ALSPAC) is a prospective, population-based pregnancy cohort study that recruited 14,541 pregnant women living in Avon, UK, with expected delivery dates between 1^st^ April 1991 and 31^st^ December 1992. Of these initial pregnancies, there were a total of 14,062 live births, and 13,988 children were alive at one year of age. The cohort has been described in detail previously (20, 21). Ethics approval for the study was obtained from the ALSPAC Ethics and Law Committee and the Local Research Ethics Committees. The study website contains details of all the data available through a fully-searchable data dictionary (http://www.bris.ac.uk/alspac/researchers/data-access/data-dictionary/).

Smoking heaviness was measured at age 15 via self-report questionnaire administered during attendance at a clinic session. Participants were asked ‘how many times have you smoked a cigarette in your lifetime?’ with the options never, 1-4, 5-20, 21-60, 61-100 and 101+ times.

At age 15, participants were administered the Vocabulary and Matrix Reasoning subsections of the Wechsler Abbreviated Scale of Intelligence (WASI) (22). General cognitive ability was calculated for each individual, adjusted for age. Scores were rescaled around the complete-case sample to a mean of 100 and a standard deviation of 15. General cognitive ability was also measured via the Wechsler Intelligence Scale for Children 3^rd^ Edition (23) at clinic at age 8.

In England, children attending state-maintained schools are educated in line with the National Curriculum, which is split in to a series of ‘Key Stages’ assessed by compulsory teacher assessments of national tests (www.gov.uk/national-curriculum/overview). Data linkage between ALSPAC and the National Pupil Database (a central repository for pupil-level educational data in England) provided educational assessment data for participants who attended state-funded school at Key Stage 4 (age 16). Data linkage was performed by a third-party company and checked by the ALSPAC team (for further information, see http://www.bristol.ac.uk/alspac/researchers/our-data/linkage/). Educational attainment at the age of 16 was quantified using a standard capped scoring method (see http://nationalpupildatabase.wikispaces.com/KS4) in which grades achieved at General Certificate of Secondary Education (GCSE) or equivalent for their best eight subjects were converted to a numerical score (A* = 58 points … G = 16 points) and summed. Capped scores (maximum = 464) were converted to a percentage for each individual.

A number of covariates were included that may confound the association between smoking and general cognitive ability or educational attainment. These were chosen after studying previous literature and in line with Mokrysz and colleagues (24). They were conceptualised as follows. Maternal and pre-birth covariates: maternal education, offspring gender, maternal depression during pregnancy, maternal alcohol, cannabis and tobacco use during pregnancy, all assessed via maternal questionnaire during pregnancy. Childhood covariates: hyperactivity and conduct problems at age 11, assessed via the strengths and difficulties questionnaire (25); depression at age 12, assessed via the moods and feelings questionnaire (26); psychotic experiences at age 12, assessed via the PLIKS semi-structured interview (27); suspected truancy at age 14 assessed via maternal questionnaire. Substance use: alcohol, cannabis and other illicit drug use measured at age 15 in the same session as the exposure.

#### Statistical analyses

Linear regression was used to assess the association between cigarette smoking at age 15 and both: a) general cognitive ability at age 15, and b) educational attainment at age 16, before and after adjustment for potential confounders. Consistent with the work of Mokrysz and colleagues (24) we excluded individuals who had maternal report of a head injury that resulted in unconsciousness due to the likelihood of impact on cognitive development (28). Furthermore, we excluded any participant who responded ‘yes’ to the question ‘have you ever used spanglers’ (a fictitious recreational drug added to the questionnaire as a quality control check). We attempted to minimise the impact of reverse causation by initially adjusting for pre-existing general cognitive ability (in the general cognitive ability analyses) or earlier educational attainment (in the education analyses) (model 2). We assessed the impact of confounding by comparing unadjusted estimates (model 1) to those adjusting for pre-birth confounders (model 3) and childhood confounders (model 4). We further adjusted for cannabis (model 5a), alcohol (5b) and illicit drugs (5c), before finally running a fully adjusted model including cannabis, alcohol and illicit drugs together (6). This approach was used to elucidate the specific impact of the covariates, given that in our previous work we have found that these variables are highly correlated with each other, and therefore that their individual impact on the strength of the association could be informative (e.g. 29). All analyses were conducted in Stata version 13 (StataCorp LP, College Station, TX). The complete case sample N for both analyses was 2,107.

#### Sensitivity Analyses

In order to investigate whether selection bias due to attrition could be impacting on the complete case analysis, multiple imputation with chained equations up to the outcome sample size was carried out for each analysis. An interaction term was included to allow us to account for earlier head injury (29). We imputed up to 100 datasets, using approximately 50 auxiliary variables to predict missing values. These included further pre-birth and childhood variables, other measures of intelligence or cognition, and earlier substance use measures. The imputed sample N was 4,954 for the general cognitive ability analysis, and 12,004 for the education analysis (as this variable was from linkage data and therefore available on many more individuals).

## Results

At age 15, 28.5% of the complete case sample of 2,107 individuals reported having smoked a cigarette, and 7.5% reported smoking more than 100 times. Table 1 shows the descriptive statistics of the covariates included in the model, stratified by heaviness of smoking. Smoking was associated with all covariates other than maternal depressive symptoms.

**Table 1.** Descriptive statistics for covariates by smoking heaviness group.

| Smoking | Never | 1-4 | 5-20 | 21-60 | 61-100 | 101+ | p |
| --- | --- | --- | --- | --- | --- | --- | --- |
| Sample (CCA) % (N) | 71.5 (1,507) | 6.7 (141) | 6.6 (139) | 5.1 (107) | 2.7 (56) | 7.4 (157) |  |
| Female | 48.6 (733) | 64.5 (91) | 67.6 (94) | 67.3 (72) | 55.4 (31) | 61.2 (96) | <0.001 |
| No higher maternal education | 13.6 (205) | 17.0 (24) | 17.3 (24) | 15.0 (16) | 21.4 (12) | 24.2 (38) | <0.001 |
| Smoking during pregnancy | 10.4 (156) | 18.4 (26) | 19.4 (27) | 15.0 (16) | 12.5 (7) | 28.7 (45) | <0.001 |
| Weekly alcohol use during pregnancy | 12.1 (182) | 22.0 (31) | 21.6 (30) | 11.2 (12) | 21.4 (12) | 15.9 (25) | 0.02 |
| Cannabis use during pregnancy | 1.0 (15) | 2.8 (4) | 3.6 (5) | 5.6 (6) | 0 | 2.6 (4) | 0.01 |
| Truancy from school (age 14) | 0.7 (10) | 0.7 (1) | 2.2 (3) | 3.7 (4) | 1.8 (1) | 8.3 (13) | <0.001 |
| Hyperactivity >3 (age 11) | 39.8 (600) | 47.5 (67) | 38.1 (53) | 54.2 (58) | 50.0 (28) | 59.9 (94) | <0.001 |
| Conduct problems >3 (age 11) | 11.0 (165) | 14.9 (21) | 13.7 (19) | 21.5 (23) | 25.0 (23) | 22.3 (35) | <0.001 |
| Psychotic-like experiences (age 12) | 10.0 (150) | 9.9 (14) | 13.0 (18) | 12.2 (13) | 19.6 (11) | 18.5 (29) | <0.001 |
| Lifetime cannabis use >20 times (age 15) | 1.0 (15) | 2.1 (3) | 7.2 (10) | 10.3 (11) | 32.1 (18) | 47.1 (74) | <0.001 |
| Lifetime alcohol use >20 times (age 15) | 24.6 (370) | 49.7 (70) | 63.3 (88) | 72.0 (77) | 87.5 (49) | 88.5 (139) | <0.001 |
| Illicit drug use since 15<sup>th</sup> birthday | 6.1 (92) | 14.9 (21) | 28.1 (39) | 33.6 (36) | 35.7 (20) | 56.1 (88) | <0.001 |
| Mean (SD) |  |  |  |  |  |  |  |
| Maternal depressive symptoms | 3.28 (2.23) | 3.52 (2.25) | 3.71 (2.26) | 3.80 (2.13) | 3.83 (2.93) | 3.70 (2.16) | 0.221 |
| Depressive symptoms (age 12) | 3.86 (1.80) | 3.96 (1.76) | 4.22 (1.80) | 4.36 (1.76) | 4.41 (1.94) | 4.54 (1.77) | <0.001 |
| General cognitive ability (age 8) | 100.7 (15.3) | 96.2 (14.2) | 100.5 (13.6) | 98.2 (15.3) | 96.6 (16.1) | 96.5 (13.8) | <0.001 |
| Educational performance (age 10/11) | 74.0 (12.6) | 69.1 (13.3) | 72.3 (12.0) | 70.3 (13.8) | 68.9 (14.0) | 68.9 (13.0) | <0.001 |

CCA: Complete case analysis.

### General cognitive ability

Heaviness of smoking at age 15 was associated with a reduction in general cognitive ability of 1.4 points per increase in category of smoking heaviness (95% CI −1.77 to −0.95), in an unadjusted analysis. After adjustment for earlier general cognitive ability, the magnitude of this association attenuated by over a third, although evidence of an association remained strong. Further adjustment for pre-birth and childhood confounders attenuated the association slightly further, but again strong evidence of an association remained. The addition of cannabis, alcohol or other illicit drugs slightly increased (model 5a and 5b) or decreased (model 5c) the size of the association. The fully adjusted model suggested a reduction in general cognitive ability of 0.7 points per increase in category of smoking heaviness (95% CI −1.24 to −0.23). The results are shown in Table 2.

**Table 2.** Linear regression analysis of smoking heaviness at age 15 and general cognitive ability at age 15, before and after adjustment for potential confounders (N=2,107).

| Model | Beta coefficient | 95% CI | P value |
| --- | --- | --- | --- |
| 1 | -1.362 | -1.773, -0.950 | <0.001 |
| 2 | -0.882 | -1.220, -0.544 | <0.001 |
| 3 | -0.721 | -1.059, -0.384 | <0.001 |
| 4 | -0.674 | -1.021, -0.327 | <0.001 |
| 5a | -0.728 | -1.199, -0.257 | 0.002 |
| 5b | -0.722 | -1.139, -0.304 | 0.001 |
| 5c | -0.618 | -1.007, -0.229 | 0.002 |
| 6 | -0.736 | -1.238, -0.233 | 0.004 |

Model 1 – Standardized general cognitive score at age 15 by unit increase of 5-level smoking heaviness category at age 15; Model 2 – as model 1 with additional adjustment for general cognitive ability at age 8; Model 3 – as model 2 with additional adjustment for pre-birth confounders (gender, maternal education, maternal smoking, alcohol and cannabis use during pregnancy, maternal depression); Model 4 – as model 3 with additional adjustment for childhood confounders (truancy, hyperactivity, conduct problems, psychotic-like experiences, depression); Model 5a – as model 4 with additional adjustment for cannabis use at age 15; Model 5b – as model 4 with additional adjustment for alcohol use at age 15; Model 5c – as model 4 with additional adjustment for illicit drug use at age 15; Model 6 – as model 4 with additional adjustment for cannabis, alcohol and other illicit drug use at age 15.

### Educational attainment

Heaviness of smoking was associated with a 2.5 percentage point decrease in educational attainment score per increase in category of smoking heaviness in the unadjusted analysis (95% CI −2.87 to −2.17). As for associations with general cognitive ability, the effect size attenuated substantially after adjustment for earlier educational attainment. Adjustment for childhood covariates and substance use further attenuated the associations, although strong evidence of an association remained. The fully adjusted model (model 6) indicated a 1.3 percentage point decrease in education score associated with each increase in smoking heaviness category (95% CI −1.60 to −0.91). These results are shown in Table 3.

**Table 3.** Linear regression of smoking heaviness at age 15 and educational attainment at age 16, before and after adjustment for potential confounders (N=2,017).

| Model | Beta coefficient | 95% CI | P value |
| --- | --- | --- | --- |
| 1 | -2.517 | -2.868, -2.166 | <0.001 |
| 2 | -1.725 | -1.969, -1.482 | <0.001 |
| 3 | -1.749 | -1.986, -1.512 | <0.001 |
| 4 | -1.561 | -1.800, -1.321 | <0.001 |
| 5a | -1.322 | -1.645, -0.999 | <0.001 |
| 5b | -1.464 | -1.751, -1.176 | <0.001 |
| 5c | -1.396 | -1.664, -1.129 | <0.001 |
| 6 | -1.254 | -1.597, -0.911 | <0.001 |

Model 1 – Educational performance score (as % of total possible) at age 16 by unit increase of 5-level smoking heaviness category at age 15; Model 2 – as model 1 with additional adjustment for educational attainment at age 11; Model 3 – as model 2 with additional adjustment for pre-birth confounders (gender, maternal education, maternal smoking, alcohol and cannabis use during pregnancy, maternal depression); Model 4 – as model 3 with additional adjustment for childhood confounders (truancy, hyperactivity, conduct problems, psychotic-like experiences, depression); Model 5a – as model 4 with additional adjustment for cannabis use at age 15; Model 5b – as model 4 with additional adjustment for alcohol use at age 15; Model 5c – as model 4 with additional adjustment for illicit drug use at age 15 Model 6 – as model 4 with additional adjustment for cannabis, alcohol and other illicit drug use at age 15.

### Multiple imputation

In the 100 imputed datasets, unadjusted associations between cigarette use and cognitive ability were somewhat smaller in magnitude than those seen in the complete case analysis (effect estimate −1.17, 95% CI −1.43 to −0.92). Adjustment for pre-birth and childhood confounders led to further attenuation. There was a small attenuation after further adjustment for cannabis, alcohol or other illicit drug use. Although effect sizes were smaller, the confidence intervals were consistent with those from the complete case analysis.

Conversely, the associations between cigarette use and educational attainment were larger in the imputed analyses than the complete case analysis (effect estimate −4.69, 95% CI −5.05 to −4.34). The pattern of attenuation was similar to that seen in the complete case analyses, although for the most part confidence intervals did not overlap with the complete case analyses. These results are shown in Supplementary Tables 1 and 2.

## Discussion

Our results indicate that smoking cigarettes at age 15 was associated with reduced cognitive ability (i.e., vocabulary and reasoning) as measured at age 15 by the WASI, and with poorer educational attainment (i.e., grades in key stage 4 national tests) at age 16. In an attempt to minimize the possibility that our results were due to cognitive ability and educational attainment making individuals more likely to smoke, we adjusted our analyses for pre-existing general cognitive ability and earlier educational attainment. This attenuated the observed associations, although strong evidence of an association remained. Although it is important to note that this pattern of attenuation is also consistent with a null causal effect and measurement error being present (30).

Associations also remained when adjusted for substance use, although we did observe some attenuation consistent with the literature suggesting that substance use is associated with poor cognitive performance and educational attainment (13). For example, cannabis is related to poorer overall learning and impaired memory (31-33). Likewise, alcohol use has been reported to be negatively associated with educational attainment and to long-lasting changes in the adult brain (13, 34). Multiple imputation of missing data allowed us to predict missing variables and, consequently, to include a larger number of ALSPAC subjects in the analysis. The results of imputation analyses for cognitive ability were broadly consistent with those seen in the complete case analysis although the strength of the associations was attenuated. Conversely, the coefficients from the imputed analyses for educational attainment were much larger than for the complete case analysis, without overlapping confidence intervals, although the pattern of attenuation after adjustment for covariates was comparable. It is possible that this is due to the larger proportion of data being imputed or due to the larger sample size available for educational attainment compared to cognitive ability.

## Study Two

### Methods

#### Data sources

We conducted two-sample MR of two smoking phenotypes (smoking initiation and lifetime smoking) on cognitive ability and educational attainment. For the smoking initiation instrument, we used summary data for the 378 independent genome-wide significant SNPs identified by the GSCAN consortium GWAS (35). For lifetime smoking (combined smoking duration, cessation and heaviness), we used summary statistics reported by Wootton and colleagues (36). The instrument consisted of 126 independent SNPs. For cognitive ability we used a GWAS of fluid intelligence conducted in the UK Biobank by the Neale lab (http://www.nealelab.is/uk-biobank/). For the smoking initiation analysis we used betas from the GSCAN GWAS with UK Biobank removed to avoid sample overlap. This was not possible for the lifetime smoking GWAS so this analysis was not conducted. For educational attainment, we used the second GWAS from the SSGAC consortium (37) discovery sample. This GWAS was chosen rather than a more recently published GWAS (38) as there was no sample overlap between exposure and outcome samples (the more recent GWAS uses Biobank data, as does the smoking GWAS). Educational attainment was defined as number of completed years of schooling.

#### Statistical analysis

We conducted four methods of Mendelian randomisation: inverse-variance weighted, MR Egger (39), weighted median (40), weighted mode (41). Each makes different assumptions about the presence of pleiotropy. A consistent direction of effect across all four methods provides the strongest evidence for a causal effect. We further calculated the MR Egger intercept, an indicator of directional pleiotropy (39). Finally, Steiger filtering was used to confirm the hypothesised direction of effect (42). Here, each SNP is examined individually to ensure that it explains more variance in the exposure than outcome. If a SNP explains more variance in the outcome, this could indicate that the outcome actually precedes the exposure on the causal pathway. Where this occurs, all analyses are re-run with these SNPs removed. All analyses were conducted using MR Base, a package for two-sample MR (43) in R (R. Core Team., 2013).

As a sensitivity analysis, multivariable MR (MVMR) was performed to investigate the independent effects of genetic liability to ADHD and smoking on cognitive ability and educational attainment (44, 45). If genetic liability to smoking initiation includes elements of impulsivity, or other related (e.g., risk-taking) traits, simultaneously modelling genetic liability to ADHD (as a marker of impulsivity) and smoking should help to disentangle this relationship. MVMR is an extension of MR that uses genetic variants associated with multiple exposures to simultaneously model the causal effect of each exposure on the outcome (44). These analyses were performed in the ALSPAC dataset from Study One.

A further sensitivity analysis using the ALSPAC dataset investigated associations between a standardised polygenic risk score for lifetime smoking, and smoking initiation with IQ measured at age 8 using the WISC. The purpose of this was to investigate the association between genetic factors associated with smoking and the outcome of interest prior to any smoking being likely to have occurred, as a negative control to test the assumption made in the later Mendelian randomization study that the SNPs have no direct effect on education aside from via the pathway of smoking. Polygenic risk scores were calculated using PLINK ‘score’ command. Genome-wide significant SNPs were identified in the previous GWAS of lifetime smoking (Wootton et al., 2018) and smoking initiation (35) and weighted by the effect sizes found in these GWAS. The WISC continuous total score was used as the outcome.

## Results

### General cognitive ability

There was some evidence for an adverse effect of smoking initiation on cognitive ability (IVW MR −0.82, 95% CI −1.04, −0.60, P = 5.36 × 10^−13^), and the direction of effect was consistent across all methods (see Table 4). Steiger filtering indicated that 86% of the SNPs for smoking initiation explained more variance in cognitive ability than smoking phenotypes (see Table 5). After Steiger filtering, the effects of smoking initiation on cognitive ability attenuated to the null (IVW MR 0.04, 95% CI −0.29 to 0.38, P = 0.81).

**Table 4.** Two-sample Mendelian randomisation analyses of the effect of smoking on educational attainment and cognitive ability.

| **Exposure** | **Outcome** | **Method** | **N SNPs** | **Beta (95% CI)** | **P-value** |
| --- | --- | --- | --- | --- | --- |
| Smoking Initiation (GSCAN) | Cognitive ability | Inverse-Variance Weighted | 376 | -0.817 ( -1.038, -0.595) | 5.36 x 10<sup>-13</sup> |
|  |  | MR Egger | 376 | -0.469 ( -0.952, 0.013) | 0.057 |
|  |  | Weighted Median | 376 | -0.597 (-0.854, -0.340) | 5.22 x 10<sup>-06</sup> |
|  |  | Weighted Mode | 376 | -0.469 (-1.274, 0.336) | 0.254 |
| Smoking Initiation (GSCAN) | Educational Attainment | Inverse-Variance Weighted | 371 | -0.197 (-0.223, -0.171) | 1.78 x 10<sup>-49</sup> |
|  |  | MR Egger | 371 | -0.145 (-0.254, -0.035) | 0.009 |
|  |  | Weighted Median | 371 | -0.159 (-0.185, -0.132) | 1.07 x 10<sup>-31</sup> |
|  |  | Weighted Mode | 371 | -0.162 (-0.251, -0.073) | 4.15 x 10<sup>-04</sup> |
| Lifetime Smoking | Educational Attainment | Inverse-Variance Weighted | 126 | - 0.393 (-0.456, -0.330) | 1.35 x 10<sup>-34</sup> |
|  |  | MR Egger | 126 | - 0.084 (-0.333, 0.165) | 0.509 |
|  |  | Weighted Median | 126 | - 0.254 (-0.321, -0.188) | 4.84 x 10<sup>-14</sup> |
|  |  | Weighted Mode | 126 | - 0.163 (-0.334, -0.009) | 0.065 |

**Table 5.**
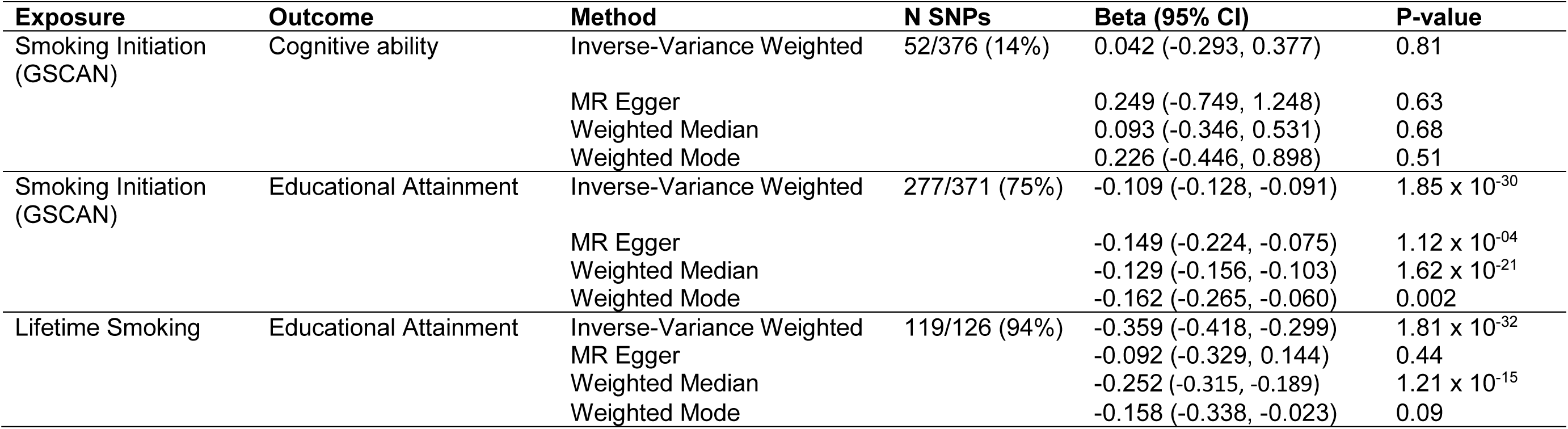
Two-sample Mendelian randomisation analyses of the effect of smoking on educational attainment and cognitive ability after Steiger filtering.

| **Exposure** | **Outcome** | **Method** | **N SNPs** | **Beta (95% CI)** | **P-value** |
| --- | --- | --- | --- | --- | --- |
| Smoking Initiation (GSCAN) | Cognitive ability | Inverse-Variance Weighted | 52/376 (14%) | 0.042 (-0.293, 0.377) | 0.81 |
|  |  | MR Egger |  | 0.249 (-0.749, 1.248) | 0.63 |
|  |  | Weighted Median |  | 0.093 (-0.346, 0.531) | 0.68 |
|  |  | Weighted Mode |  | 0.226 (-0.446, 0.898) | 0.51 |
| Smoking Initiation (GSCAN) | Educational Attainment | Inverse-Variance Weighted | 277/371 (75%) | -0.109 (-0.128, -0.091) | 1.85 x 10<sup>-30</sup> |
|  |  | MR Egger |  | -0.149 (-0.224, -0.075) | 1.12 x 10<sup>-04</sup> |
|  |  | Weighted Median |  | -0.129 (-0.156, -0.103) | 1.62 x 10<sup>-21</sup> |
|  |  | Weighted Mode |  | -0.162 (-0.265, -0.060) | 0.002 |
| Lifetime Smoking | Educational Attainment | Inverse-Variance Weighted | 119/126 (94%) | -0.359 (-0.418, -0.299) | 1.81 x 10<sup>-32</sup> |
|  |  | MR Egger |  | -0.092 (-0.329, 0.144) | 0.44 |
|  |  | Weighted Median |  | -0.252 (-0.315, -0.189) | 1.21 x 10<sup>-15</sup> |
|  |  | Weighted Mode |  | -0.158 (-0.338, -0.023) | 0.09 |

### Multivariable MR

When estimating the causal effect of both genetic liability to ADHD and smoking initiation, there was strong evidence of an association of a negative effect of genetic liability to ADHD on cognitive ability (−0.171, 95% CI −0.26 to −0.09, P = 9.14 × 10^−5^), independently of smoking initiation. There was also some evidence of an association for a negative effect of genetic liability to smoking initiation on cognitive ability (−0.405, 95% CI - 0.72 to −0.09, P = 0.012), independently of ADHD. However, there was some evidence of pleiotropy within these instruments and instrument strength was weak. As such these estimates should be interpreted with caution. These results are shown in Table 6.

**Table 6.**
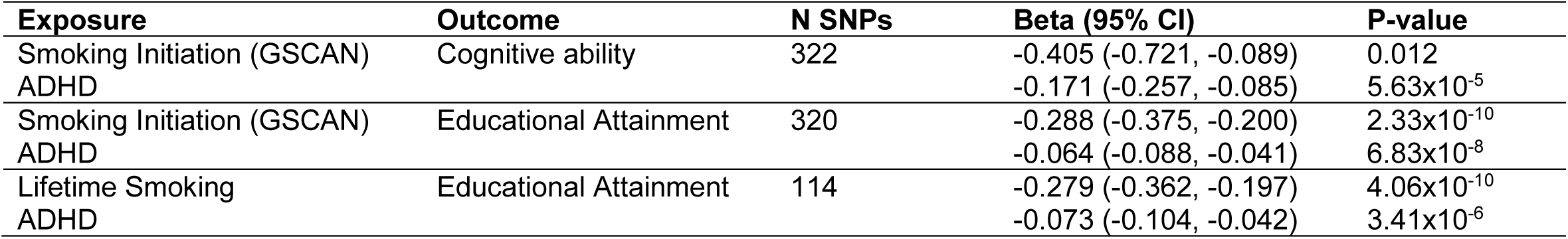
Multivariable Mendelian randomization analyses using summary statistics simultaneously modelling genetic liability to ADHD and smoking on educational attainment or cognitive ability.

### Educational attainment

Of the 126 SNPs associated with lifetime smoking (36), all were available in the GWAS of educational attainment (37) and the GWAS of cognitive ability (http://www.nealelab.is/uk-biobank/). Of the 378 SNPs associated with smoking initiation (35), 371 were available in the GWAS of educational attainment (37) and 376 were available in the GWAS of cognitive ability (http://www.nealelab.is/uk-biobank/). For both smoking phenotypes, there was a consistent adverse effect of smoking on educational attainment across all four MR methods (see Table 4). The evidence was strongest for the effect of smoking initiation on educational attainment with genetic risk for smoking initiation associated with a 0.39 SD decrease in years of schooling (95% CI −0.456 to −0.330, P = 1.35 × 10^−34^). Both MR Egger intercepts suggested weak evidence of directional pleiotropy. After Steiger filtering, 94% of SNPs were retained for the lifetime smoking analysis, and 75% of SNPs for the smoking initiation analysis (see Table 5), and the resultant analyses found strong evidence for an effect of smoking on educational attainment (e.g., for lifetime smoking, using 119 of the 126 original SNPs IVW MR −0.359, 95% CI −0.418 to −0.299, P = 1.81 × 10^−32^).

### Multivariable MR

When estimating the independent causal effects of genetic liability to ADHD and smoking initiation, there was strong evidence of a negative effect of genetic liability to ADHD on educational attainment (−0.064, 95% CI: −0.088, −0.041, p=6.86×10^−8^). We also found strong evidence of a negative effect of genetic liability to smoking initiation on educational attainment (−0.288, 95% CI −0.375 to −0.20, P = 2.33 × 10^−10^). Similar effects were found when looking at ADHD and lifetime smoking, with strong evidence of a negative effect of genetic liability to both ADHD (−0.073, 95% CI −0.104 to −0.042, P = 2.41×10^−6^) and lifetime smoking (−0.279, 95% CI −0.362 to −0.197, P = 1.93×10^−11^) on educational attainment. These results are shown in Table 6. However, in both analyses, there was evidence of pleiotropy and instrument strength was weak (Supplementary Table 3).

### Sensitivity analyses

In the ALSPAC dataset associations were found between the polygenic risk scores for both lifetime smoking and smoking initiation, with IQ measured at age 8 years. This was true in the whole sample (N=5,300), and when restricted to individuals who reported never having tried a cigarette (N=4,650). These results are shown in Table 7. The multivariable MR examining smoking and ADHD on IQ at age 15 was also run in the ALSPAC sample, although this was very underpowered (see Supplementary Table 4).

**Table 7.** Associations between standardised polygenic risk scores for lifetime smoking (LS) and smoking initiation (SI) with IQ (measured as total score on the WISC) at age 8 in the ALSPAC offspring.

|  |  | **Beta** | **95% CI** | **P-value** | **N** |
| --- | --- | --- | --- | --- | --- |
| Whole sample | LS | -0.934 | -1.37, -0.50 | $2.8 \times 10^{-5}$ | 5,300 |
| | SI | -0.749 | -1.17, -0.30 | $9.0 \times 10^{-4}$ | 5,300 |
| Restricted to individuals who report never trying a cigarette | LS | -0.893 | -1.34, -0.44 | $1.0 \times 10^{-4}$ | 4,950 |
| | SI | -0.785 | -1.24, -0.33 | $6.3 \times 10^{-4}$ | 4,950 |

Analyses are adjusted for sex, age, and first 5 principal components

## Discussion

These results broadly replicate the findings from our observational analyses in Study One, and provide some evidence for a causal effect of smoking on education. The results for cognitive ability are more mixed, but may indicate common genetic influence on both cognition and smoking. Interestingly, there was stronger evidence for pleiotropy in the analysis of lifetime smoking on educational attainment, compared with the analysis of smoking initiation on educational attainment. Given that educational attainment is largely realised by early adulthood, smoking initiation may be the more appropriate exposure to examine (unlike for most outcomes, where the effects of smoking develop over a lifetime of exposure). We have also previously shown that educational attainment impacts on smoking (3, 4, 46), with these effects likely to continue to play out over the lifetime (e.g., via effects on heaviness of smoking and smoking cessation). Interestingly, multivariable MR analyses suggest that there is no independent effect of general cognitive ability on smoking when the effect of educational attainment is taken into account (4).

Taken together, these results and our previous findings indicate a complex, bidirectional causal relationship between smoking and educational attainment. Evidence for the causal effect of smoking on cognition is more mixed. This is perhaps to be expected, as cognition is less susceptible to the impact of the environment than educational attainment is likely to be (47). It is also important to bear in mind that MR analyses between lifetime smoking and cognitive ability were not possible due to the use of UK Biobank data in both the exposure and outcome GWAS. The implementation of Steiger filtering and subsequent removal of 86% of the 376 initial SNPs may indicate evidence of reverse causality or a common biological cause, although power is reduced for these analyses due to the removal of identified SNPs. Critically, the results of our negative control analyses, which indicate that the genetic instruments for smoking are associated with general cognitive ability before individuals have begun smoking, suggests direct effects of these variants on cognition. One possibility is that these variants capture a common mechanism (e.g., trait impulsivity) that has effects on both smoking and cognition, and we explored this using the MVMR analysis.

## General Discussion

Our study provides observational evidence that smoking is associated with reduced general cognitive ability and educational attainment (Study One), and MR evidence that the associations with educational attainment are likely to be causal (Study Two). These results provide further support for the importance of preventive interventions in schools to reduce the uptake of smoking in young people. The triangulation of results across different methods, each with their own strengths, limitations and sources of bias, is an important strength of our approach, and allows us to be more confident that our results reflect genuine causal relationships, and are robust.

There are a number of limitations to this study that should be considered. First, smoking was based on self-report as there was no objective measure available in either our observational or MR analyses. Second, in our observational analyses, the WASI was not used in its entirety and therefore the estimated score was less reliable than a full IQ score (Mokrysz et al., 2016). Third, our observational analyses are cross sectional, meaning reverse causality cannot be ruled out, although we attempt to minimise any impact by adjusting for earlier cognitive ability or educational attainment. Similarly, we cannot exclude the possibility that a common risk factor is operating that influences both smoking and lower cognitive abilities and educational attainment. However, the results from our MR analyses protect against this and allow us to triangulate the results of both approaches to suggest that smoking does indeed causally influence educational attainment. Indeed, the results of our negative control analyses in ALSPAC provide some evidence for this possibility.

MR should in principle allow us to assess the associations between smoking and education without the confounding from socio-economic status, if the assumptions of MR have been met. However, if individuals who smoke are more likely to reproduce with other individuals who smoke, as some studies have indicated (48), the assumptions made by MR may not hold true. Given the social patterning of smoking behaviour, cigarette use might be a marker for many aspects of deprivation that would likely impact on years of education (for example the need to leave school to begin employment), an interpretation that cannot be ruled out by these data. Relatedly, it may be the case that the questions used are capturing different phenotypes to those intended. This may be the case for smoking initiation – asking an individual whether they have ever tried smoking may be a strong predictor of traits such as impulsivity, rather than smoking itself. However, there is evidence from other studies using different designs that do not have these limitations that smoking might causally impact on educational attainment, including instrumental variable analyses in different cultural settings (49), allowing for further triangulation of results.

It has long been suggested that nicotine may be a cognitive enhancer, which is apparently paradoxical given our results. A number of studies have found that acute nicotine intoxication can transiently improve cognitive abilities (50). However, the acute effects of a substance may differ from the longer-term impact of regular use. For example, regular cigarette users often report that smoking will reduce feelings of anxiety, while regular smoking is predictive of increased subsequent anxiety (51). Nicotine withdrawal may be responsible for both these seemingly contradictory effects – the removal of nicotine withdrawal symptoms could appear to improve cognitive ability and mood, by reversing impairment caused by withdrawal symptoms (50, 52). It is also interesting to note the difference between long-term smoking and the impact of acute nicotine in the lab, which is often delivered via routes other than smoking (53). The mechanisms by which cigarette use might impact on educational attainment are harder to elucidate. There is some evidence to suggest that smoking may lead to adverse mental health outcomes, or externalising behaviours (54). It is possible that these, in turn, could influence educational outcomes and/or cognitive ability, although our data cannot interrogate this hypothesis directly.

We have used two different methodologies to investigate the link between smoking and education and cognitive ability. Results from both studies suggest that smoking can affect educational attainment as early in life as adolescence. While both methods used have limitations, they have different limitations, and therefore the consistency across both methods provides further evidence that this is a true effect, rather than a product of bias or confounding. Future research should strive to ascertain the mechanisms that underpin this association, which could inform early interventions aimed at preventing smoking uptake, for example within the school environment, as these could support higher educational attainment and, ultimately, reduce the health and social inequality that derives from both smoking and low education.

## Data Availability

Data used is from the ALSPAC cohort (access details available at http://www.bristol.ac.uk/alspac/researchers/access/), and from publicly available published GWAS summary statistics.

## Acknowledgements

We are extremely grateful to all the families who took part in this study, the midwives for their help in recruiting them, and the whole ALSPAC team, which includes interviewers, computer and laboratory technicians, clerical workers, research scientists, volunteers, managers, receptionists and nurses. The UK Medical Research Council and Wellcome Trust (Grant ref: 102215/2/13/2) and the University of Bristol provide core support for ALSPAC. MRM is a member of the UK Centre for Tobacco Control Studies, a UKCRC Public Health Research: Centre of Excellence. Funding from British Heart Foundation, Cancer Research UK, Economic and Social Research Council, Medical Research Council, and the National Institute for Health Research, under the auspices of the UK Clinical Research Collaboration, is gratefully acknowledged. This work was supported by the Medical Research Council (MC_UU_00011/7), and the NIHR Biomedical Research Centre at the University Hospitals Bristol NHS Foundation Trust and the University of Bristol. The views expressed in this publication are those of the authors and not necessarily those of the NHS, the National Institute for Health Research or the Department of Health and Social Care. The funders were not involved in any aspect of the study design, collection, analysis, interpretation of data, the writing of the report, or the submission to the journal.

## References

1. Jefferis BJ, Power C, Graham H, Manor O. Changing social gradients in cigarette smoking and cessation over two decades of adult follow-up in a British birth cohort. Journal of public health. 2004;26(1):13–8.

2. Peretti-Watel P, Seror V, Constance J, Beck F. Poverty as a smoking trap. The International journal on drug policy. 2009;20(3):230–6.

3. Gage SH, Bowden J, Davey Smith G, Munafo MR. Investigating causality in associations between education and smoking: a two-sample Mendelian randomization study. International journal of epidemiology. 2018.

4. Sanderson E, Davey Smith G, Bowden J, Munafo MR. Mendelian randomisation analysis of the effect of educational attainment and cognitive ability on smoking behaviour. Nature communications. 2019;10(1):2949.

5. Chassin L, Presson CC, Pitts SC, Sherman SJ. The natural history of cigarette smoking from adolescence to adulthood in a midwestern community sample: multiple trajectories and their psychosocial correlates. Health Psychol. 2000;19(3):223–31.

6. Ellickson PL, Tucker JS, Klein DJ. High-risk behaviors associated with early smoking: results from a 5-year follow-up. J Adolesc Health. 2001;28(6):465–73.

7. Orlando M, Tucker JS, Ellickson PL, Klein DJ. Developmental trajectories of cigarette smoking and their correlates from early adolescence to young adulthood. J Consult Clin Psychol. 2004;72(3):400–10.

8. Orpinas P, Lacy B, Nahapetyan L, Dube SR, Song X. Cigarette Smoking Trajectories From Sixth to Twelfth Grade: Associated Substance Use and High School Dropout. Nicotine & tobacco research : official journal of the Society for Research on Nicotine and Tobacco. 2016;18(2):156–62.

9. Fergusson DM, Horwood LJ, Ridder EM. Show me the child at seven II: Childhood intelligence and later outcomes in adolescence and young adulthood. Journal of child psychology and psychiatry, and allied disciplines. 2005;46(8):850–8.

10. Latvala A, Rose RJ, Pulkkinen L, Dick DM, Korhonen T, Kaprio J. Drinking, smoking, and educational achievement: cross-lagged associations from adolescence to adulthood. Drug and alcohol dependence. 2014;137:106–13.

11. Maralani V. Understanding the links between education and smoking. Soc Sci Res. 2014;48:20–34.

12. Beman DS. Risk factors leading to adolescent substance abuse. Adolescence. 1995;30(117):201–8.

13. Cox RG, Zhang L, Johnson WD, Bender DR. Academic performance and substance use: findings from a state survey of public high school students. J Sch Health. 2007;77(3):109–15.

14. Brunswick AF, Messeri PA. Origins of cigarette smoking in academic achievement, stress and social expectations: Does gender make a difference? The Journal of Early Adolescence. 1984;4(4):353–70.

15. Davey Smith G, Hemani G. Mendelian randomization: genetic anchors for causal inference in epidemiological studies. Human molecular genetics. 2014;23(R1):R89–98.

16. Davey Smith G, Ebrahim S. ‘Mendelian randomization’: can genetic epidemiology contribute to understanding environmental determinants of disease? International journal of epidemiology. 2003;32(1):1–22.

17. Gage SH, Munafo MR, Davey Smith G. Causal Inference in Developmental Origins of Health and Disease (DOHaD) Research. Annual review of psychology. 2016;67:567–85.

18. Lawlor DA, Tilling K, Davey Smith G. Triangulation in aetiological epidemiology. Int J Epidemiol. 2016;45(6):1866–86.

19. Munafo MR, Davey Smith G. Robust research needs many lines of evidence. Nature. 2018;553(7689):399–401.

20. Boyd A, Golding J, Macleod J, Lawlor DA, Fraser A, Henderson J, et al. Cohort Profile: the ‘children of the 90s’--the index offspring of the Avon Longitudinal Study of Parents and Children. International journal of epidemiology. 2013;42(1):111–27.

21. Fraser A, Macdonald-Wallis C, Tilling K, Boyd A, Golding J, Davey Smith G, et al. Cohort Profile: the Avon Longitudinal Study of Parents and Children: ALSPAC mothers cohort. International journal of epidemiology. 2013;42(1):97–110.

22. Wechsler D. Wechsler Abbreviated Scale of Intelligence. San Antonio, TX: Psychological Corporation; 1999.

23. Wechsler D, Golombok J, Rust J. Wechsler Intelligence Scale for Children Sidcup, United Kingdom: The Psychological Corporation; 1992.

24. Mokrysz C, Landy R, Gage SH, Munafo MR, Roiser JP, Curran HV. Are IQ and educational outcomes in teenagers related to their cannabis use? A prospective cohort study. J Psychopharmacol. 2016;30(2):159–68.

25. Goodman R. The Strengths and Difficulties Questionnaire: a research note. Journal of child psychology and psychiatry, and allied disciplines. 1997;38(5):581–6.

26. Costello EJ, Angold A. Scales to assess child and adolescent depression: checklists, screens, and nets. J Am Acad Child Adolesc Psychiatry. 1988;27(6):726–37.

27. Zammit S, Odd D, Horwood J, Thompson A, Thomas K, Menezes P, et al. Investigating whether adverse prenatal and perinatal events are associated with non-clinical psychotic symptoms at age 12 years in the ALSPAC birth cohort. Psychological medicine. 2009;39(9):1457–67.

28. Azouvi P, Arnould A, Dromer E, Vallat-Azouvi C. Neuropsychology of traumatic brain injury: An expert overview. Rev Neurol (Paris). 2017;173(7-8):461–72.

29. Gage SH, Hickman M, Heron J, Munafo MR, Lewis G, Macleod J, et al. Associations of cannabis and cigarette use with depression and anxiety at age 18: findings from the Avon Longitudinal Study of Parents and Children. PLoS One. 2015;10(4):e0122896.

30. Fewell Z, Davey Smith G, Sterne JA. The impact of residual and unmeasured confounding in epidemiologic studies: a simulation study. Am J Epidemiol. 2007;166(6):646–55.

31. Lovell ME, Bruno R, Johnston J, Matthews A, McGregor I, Allsop DJ, et al. Cognitive, physical, and mental health outcomes between long-term cannabis and tobacco users. Addict Behav. 2018;79:178–88.

32. Meier MH, Caspi A, Ambler A, Harrington H, Houts R, Keefe RS, et al. Persistent cannabis users show neuropsychological decline from childhood to midlife. Proc Natl Acad Sci U S A. 2012;109(40):E2657–64.

33. Schuster RM, Crane NA, Mermelstein R, Gonzalez R. Tobacco may mask poorer episodic memory among young adult cannabis users. Neuropsychology. 2015;29(5):759–66.

34. Crews FT, Vetreno RP, Broadwater MA, Robinson DL. Adolescent Alcohol Exposure Persistently Impacts Adult Neurobiology and Behavior. Pharmacol Rev. 2016;68(4):1074–109.

35. Liu M, Jiang Y, Wedow R, Li Y, Brazel DM, Chen F, et al. Association studies of up to 1.2 million individuals yield new insights into the genetic etiology of tobacco and alcohol use. Nature genetics. 2019;51(2):237–44.

36. Wootton RE, Richmond RC, Stuijfzand BG, Lawn RB, Sallis HM, Taylor GMJ, et al. Causal effects of lifetime smoking on risk for depression and schizophrenia: Evidence from a Mendelian randomisation study. BioRxiv. 2018.

37. Okbay A, Beauchamp JP, Fontana MA, Lee JJ, Pers TH, Rietveld CA, et al. Genome-wide association study identifies 74 loci associated with educational attainment. Nature. 2016;533(7604):539–42.

38. Lee JJ, Wedow R, Okbay A, Kong E, Maghzian O, Zacher M, et al. Gene discovery and polygenic prediction from a genome-wide association study of educational attainment in 1.1 million individuals. Nature genetics. 2018.

39. Bowden J, Davey Smith G, Burgess S. Mendelian randomization with invalid instruments: effect estimation and bias detection through Egger regression. International journal of epidemiology. 2015;44(2):512–25.

40. Bowden J, Davey Smith G, Haycock PC, Burgess S. Consistent Estimation in Mendelian Randomization with Some Invalid Instruments Using a Weighted Median Estimator. Genetic epidemiology. 2016;40(4):304–14.

41. Hartwig FP, Davey Smith G, Bowden J. Robust inference in summary data Mendelian randomisation via the zero modal pleiotropy assumption. Int J Epidemiol. 2017.

42. Hemani G, Tilling K, Davey Smith G. Orienting the causal relationship between imprecisely measured traits using GWAS summary data. PLoS genetics. 2017;13(11):e1007081.

43. Hemani G, Zheng J, Elsworth B, Wade KH, Haberland V, Baird D, et al. The MR-Base platform supports systematic causal inference across the human phenome. eLife. 2018;7.

44. Burgess S, Thompson SG. Multivariable Mendelian randomization: the use of pleiotropic genetic variants to estimate causal effects. Am J Epidemiol. 2015;181(4):251–60.

45. Sanderson E, Davey Smith G, Windmeijer F, Bowden J. An examination of multivariable Mendelian randomization in the single-sample and two-sample summary data settings. International journal of epidemiology. 2018.

46. Davies NM, Dickson M, Davey Smith G, van den Berg GJ, Windmeijer F. The causal effects of education on health outcomes in the UK Biobank. Nat Hum Behav. 2018;2(2):117–25.

47. Ritchie SJ, Tucker-Drob EM. How Much Does Education Improve Intelligence? A Meta-Analysis. Psychol Sci. 2018;29(8):1358–69.

48. Agrawal A, Heath AC, Grant JD, Pergadia ML, Statham DJ, Bucholz KK, et al. Assortative mating for cigarette smoking and for alcohol consumption in female Australian twins and their spouses. Behav Genet. 2006;36(4):553–66.

49. Zhao M, Konishi Y, Glewwe P. Does smoking make one dumber? Evidence from teenagers in rural China. Gansu Survey of Children and Families Papers. 2010.

50. Campos MW, Serebrisky D, Castaldelli-Maia JM. Smoking and Cognition. Current drug abuse reviews. 2016;9(2):76–9.

51. Fluharty M, Taylor AE, Grabski M, Munafo MR. The Association of Cigarette Smoking With Depression and Anxiety: A Systematic Review. Nicotine & tobacco research : official journal of the Society for Research on Nicotine and Tobacco. 2017;19(1):3–13.

52. Niemegeers P, Dumont GJ, Quisenaerts C, Morrens M, Boonzaier J, Fransen E, et al. The effects of nicotine on cognition are dependent on baseline performance. Eur Neuropsychopharmacol. 2014;24(7):1015–23.

53. Heishman SJ, Kleykamp BA, Singleton EG. Meta-analysis of the acute effects of nicotine and smoking on human performance. Psychopharmacology (Berl). 2010;210(4):453–69.

54. Gage SH, Jones HJ, Taylor AE, Burgess S, Zammit S, Munafo MR. Investigating causality in associations between smoking initiation and schizophrenia using Mendelian randomization. Scientific reports. 2017;7:40653.

